# Incidence and outcomes of post-transplant lymphoproliferative disorders in lung transplant patients: analysis of ISHLT Registry

**DOI:** 10.1101/2020.01.27.20019042

**Authors:** L. Zaffiri, A. Long, M.L. Neely, W. Cherikh, D.C. Chambers, L. D. Snyder

## Abstract

**Background:** Post-transplant lymphoproliferative disorder (PTLD) is a life-threatening complication following lung transplant. We aimed to study the incidence of and risk factors for PTLD in adult lung transplant recipients.

**Methods:** The International Society of Heart and Lung Transplant (ISHLT) Registry was used to identify adult, first-time, single and bilateral lung transplant recipients with at least one year follow-up and from centers reporting PTLD between January 2006 and June 2015. Kaplan Meier method was used to describe timing and distribution of PTLD. Univariable and multivariable Cox proportional hazards regression models were used to examine the clinical characteristics associated with PTLD.

**Results:** Of the 19,309 lung transplant recipients in the analysis cohort, we identified 454 cases of PTLD. Cumulative incidence of PTLD was 1.1% (95% CI = 1.0%, 1.3%) at 1 year and 4.1% (95% CI= 3.6%, 4.6%) at 10-years. We observed that 47.4% of all PTLD cases occurred within the first year following lung transplantation. In the multivariable model, independent risk factors for PTLD included age, EBV mismatch and native lung diseases. The risk of PTLD during the first year after transplant increased with increasing age in patients between 45 to 62 years at time of transplantation; the inverse was true for ages less than 45 year or greater than 62 years. Finally, receiving a donor organ with human leukocyte antigen (HLA) types A1 and A24 was associated with an increased risk of PTLD while the recipient HLA type DR11 was associated with a decreased risk.

**Conclusions:** Our study indicates that PTLD is a relatively rare complication among adult lung transplant recipients. We identified clinical characteristics that are associated with increased risk of PTLD.

## INTRODUCTION

Post-transplant lymphoproliferative disorder (PTLD) represents a serious complication following solid organ transplantation (SOT) and is associated with significant morbidity and mortality^1^. After skin cancer, PTLD is the second most common malignancy following transplantation^2,3^. The risk of developing PTLD after SOT is estimated as 20-fold higher than the general population ^4^. The highest incidence of PTLD has been described in small intestine transplant recipients followed by lung and heart-lung recipients. ^2,5-7^. However, given the relative rare occurrence of PTLD, there are few single-center studies analyzing incidence and risk factors associated with PTLD in adult lung transplant recipients. Most PTLD cases occur during the first year, however late cases have been reported 5-7 years after transplant ^8-10^. Risk factors associated with PTLD in lung transplant recipients include: EBV infection and EBV mismatch in EBV seronegative recipients, cytomegalovirus (CMV) infection, cystic fibrosis (CF), more aggressive immunosuppression and recurrent episodes of rejection ^9,11-13^. Less clear are the risk factors associated with late cases of PTLD ^14^. However, older age and differences in morphology and location suggest that other factors might play a role in the development of late cases ^15-17^.

Another possible association has been described between human leukocyte antigen (HLA) and PTLD ^18,19^. Donor or recipient HLAs could modulate EBV-specific T cell responses and influence PTLD risk. Several contradictory findings have been reported ^18,20-22^, however a dedicated analysis of this association in lung transplant recipients has not been published.

The aims of the present study were to analyze the International Society for Heart and Lung transplant (ISHLT) Thoracic Organ Transplant Registry in order to describe the incidence of PTLD and identify clinical characteristics associated with PTLD in adult lung transplant recipients.

## METHODS

### Study design

We analyzed the ISHLT Registry, an international longitudinal voluntary database that incorporates country and consortium transplant data reported by individual centers. Our retrospective analysis included 19,309 lung transplants performed between January 2006 and June 2015 who survived at least 1-year after transplant.

### Primary end-points and covariables

The primary end-point of interest was time to development of PTLD after lung transplantation. Outcomes were evaluated at the time of yearly follow-up. Primary end-point was censored at last follow-up, at patient death, or at retransplant (i.e., graft failure). We considered donor variables and recipient pre- and peri-transplant variables based on clinical relevance and reported associations with PTLD from prior studies. All covariables considered in the association analysis are listed in table 1.

**Table 1.** Summary of pre- and peri-transplant characteristics.

|  | Overall<br>(N=19309) |  | Subjects with PTLD<br>(N=454) |  | Subjects without PTLD<br>(N=18855) |  |
| --- | --- | --- | --- | --- | --- | --- |
| Patient Characteristics * | Summary Measure | Missing | Summary Measure | Missing | Summary Measure | Missing |
| Recipient Age at Transplant (years) | 57 (47, 63) | 0 (0.00%) | 58 (45, 62) | 0 (0.00%) | 57 (47, 63) | 0 (0.00%) |
| Transplant Procedure Type |  | 0 (0.00%) |  | 0 (0.00%) |  | 0 (0.00%) |
| Single | 7012 (36.31%) |  | 153 (33.70%) |  | 6859 (36.38%) |  |
| Bilateral | 12297 (63.69%) |  | 301 (66.30%) |  | 11996 (63.62%) |  |
| Female | 8320 (43.09%) | 0 (0.00%) | 173 (38.11%) | 0 (0.00%) | 8147 (43.21%) | 0 (0.00%) |
| Total Ischemia Time Left and Right Lung (hours) | 4.9 (3.9, 6) | 1101 (5.70%) | 4.92 (3.9, 6.2) | 40 (8.81%) | 4.9 (3.9, 6) | 1061 (5.63%) |
| ABO Blood Group |  | 1 (0.01%) |  | 0 (0.00%) |  | 1 (0.01%) |
| A | 7785 (40.32%) |  | 191 (42.07%) |  | 7594 (40.28%) |  |
| B | 2147 (11.12%) |  | 56 (12.33%) |  | 2091 (11.09%) |  |
| AB | 747 (3.87%) |  | 18 (3.96%) |  | 729 (3.87%) |  |
| O | 8629 (44.69%) |  | 189 (41.63%) |  | 8440 (44.76%) |  |
| Donor/Recipient ABO Match | 17813 (92.25%) | 1 (0.01%) | 422 (92.95%) | 0 (0.00%) | 17391 (92.24%) | 1 (0.01%) |
| Native Lung Disease |  | 10 (0.05%) |  | 0 (0.00%) |  | 10 (0.05%) |
| Cystic Fibrosis | 2633 (13.64%) |  | 90 (19.82%) |  | 2543 (13.49%) |  |
| Obstructive | 7707 (39.91%) |  | 146 (32.16%) |  | 7561 (40.10%) |  |
| Restrictive | 8338 (43.18%) |  | 204 (44.93%) |  | 8134 (43.14%) |  |
| Vascular | 621 (3.22%) |  | 14 (3.08%) |  | 607 (3.22%) |  |
| Functional Status at Time of Transplant |  | 821 (4.25%) |  | 39 (8.59%) |  | 782 (4.15%) |
| Performs Activities of Daily Living with No Assistance | 2683 (13.90%) |  | 82 (18.06%) |  | 2601 (13.79%) |  |

|  | Overall<br>(N=19309) |  | Subjects with PTLT<br>(N=454) |  | Subjects without PTLT<br>(N=18855) |  |
| --- | --- | --- | --- | --- | --- | --- |
| Patient Characteristics* | Summary Measure | Missing | Summary Measure | Missing | Summary Measure | Missing |
| Performs Activities of Daily Living with Some Assistance | 10872 (56.31%) |  | 228 (50.22%) |  | 10644 (56.45%) |  |
| Performs Activities of Daily Living with Total Assistance | 4933 (25.55%) |  | 105 (23.13%) |  | 4828 (25.61%) |  |
| Serum Albumin Prior to Transplant | 4 (3.6, 4.3) | 3745 (19.40%) | 4 (3.6, 4.2) | 109 (24.01%) | 4 (3.6, 4.3) | 3636 (19.28%) |
| History of Prior Malignancy | 1236 (6.40%) | 218 (1.13%) | 24 (5.29%) | 13 (2.86%) | 1212 (6.43%) | 205 (1.09%) |
| PRA% - Class 1 > 0 | 2964 (15.35%) | 5785 (29.96%) | 51 (11.23%) | 184 (40.53%) | 2913 (15.45%) | 5601 (29.71%) |
| PRA% - Class 2 > 0 | 2036 (10.54%) | 6885 (35.66%) | 30 (6.61%) | 211 (46.48%) | 2006 (10.64%) | 6674 (35.40%) |
| Recipient EBV Positive | 14750 (76.39%) | 2392 (12.39%) | 242 (53.30%) | 66 (14.54%) | 14508 (76.95%) | 2326 (12.34%) |
| Donor EBV Positive | 11499 (59.55%) | 6775 (35.09%) | 223 (49.12%) | 202 (44.49%) | 11276 (59.80%) | 6573 (34.86%) |
| Donor/Recipient EBV Match |  | 3234 (16.75%) |  | 122 (26.87%) |  | 3112 (16.50%) |
| Donor Positive/Recipient Negative | 1220 (6.32%) |  | 82 (18.06%) |  | 1138 (6.04%) |  |
| Donor Negative/Recipient Negative | 105 (0.54%) |  | 8 (1.76%) |  | 97 (0.51%) |  |
| Recipient Positive | 14750 (76.39%) |  | 242 (53.30%) |  | 14508 (76.95%) |  |
| Recipient CMV Positive | 10189 (52.77%) | 1861 (9.64%) | 202 (44.49%) | 51 (11.23%) | 9987 (52.97%) | 1810 (9.60%) |
| Donor CMV Positive | 11697 (60.58%) | 142 (0.74%) | 247 (54.41%) | 10 (2.20%) | 11450 (60.73%) | 132 (0.70%) |
| Donor/Recipient CMV Match |  | 1895 (9.81%) |  | 51 (11.23%) |  | 1844 (9.78%) |
| Donor Positive/Recipient Negative | 4289 (22.21%) |  | 113 (24.89%) |  | 4176 (22.15%) |  |
| Donor Negative/Recipient Negative | 2936 (15.21%) |  | 88 (19.38%) |  | 2848 (15.10%) |  |
| Recipient Positive | 10189 (52.77%) |  | 202 (44.49%) |  | 9987 (52.97%) |  |
| Recipient HBV Positive | 1002 (5.19%) | 1160 (6.01%) | 18 (3.96%) | 46 (10.13%) | 984 (5.22%) | 1114 (5.91%) |
| Donor HBV Positive | 477 (2.47%) | 80 (0.41%) | 10 (2.20%) | 10 (2.20%) | 467 (2.48%) | 70 (0.37%) |
| Recipient HCV Positive | 317 (1.64%) | 1812 (9.38%) | 7 (1.54%) | 54 (11.89%) | 310 (1.64%) | 1758 (9.32%) |

**Table 1. Summary of pre- and peri-transplant characteristics**
|  | Overall<br>(N=19309) |  | Subjects with PTLD<br>(N=454) |  | Subjects without PTLD<br>(N=18855) |  |
| --- | --- | --- | --- | --- | --- | --- |
| Patient Characteristics* | Summary Measure | Missing | Summary Measure | Missing | Summary Measure | Missing |
| Donor HCV Positive | 14 (0.07%) | 121 (0.63%) | 0 (0.00%) | 18 (3.96%) | 14 (0.07%) | 103 (0.55%) |
| Donor History of High Risk Behaviors | 1526 (7.90%) | 3887 (20.13%) | 37 (8.15%) | 142 (31.28%) | 1489 (7.90%) | 3745 (19.86%) |
| Donor Age (years) | 32 (21, 46) | 1 (0.01%) | 29 (21, 44) | 0 (0.00%) | 32 (21, 46) | 1 (0.01%) |
| Donor History of Cancer | 354 (1.83%) | 263 (1.36%) | 3 (0.66%) | 12 (2.64%) | 351 (1.86%) | 251 (1.33%) |
| Donor Steroids | 14406 (74.61%) | 900 (4.66%) | 339 (74.67%) | 33 (7.27%) | 14067 (74.61%) | 867 (4.60%) |
| *Continuous variables are summarized by median (25th percentile, 75th percentile); Categorical variables are summarized by count (percentage).<br>PRA: panel reactive antibody; HBV: hepatitis B virus; HCV: hepatitis C virus |  |  |  |  |  |  |

Data collected in the ISHLT Registry is derived from real-world practice patterns reported into smaller registries. As such, missing data fields were expected based on specific registry collection instruments as well as varying completeness of collection. Covariates with high-levels of missingness (> 25%) were not considered in the inferential analysis.

### Statistical Analysis

To describe timing and distribution of PLTD events after lung transplantation, Kaplan Meier (KM) method was used to obtain cumulative incidence estimates. Univariable and multivariable Cox proportional hazards regression models were used to examine the association between time to development of PTLD and recipient and donor characteristics. Associations were considered statistically significant if p < 0.05. To assess proportional hazards assumption, interaction between log time-to-event and each covariable was tested. If log-time--covariable interaction was statistically significant, the relationship between the covariable and PTLD was characterized by the interaction p-value and by reporting a set of hazard ratios (HRs) and 95% confidence intervals over follow-up period (specifically, at 1-year, 5-years, and 10-years); otherwise, a single HR, confidence interval, and p-value were reported for the entire follow-up period.

For continuous covariables, linearity assumption was assessed by performing a lack-of-fit test comparing a linear fit with a non-linear fit based on a restricted cubic spline (RCS) with 3 knots. If non-linearity was statistically significant, a piecewise linear spline was used to account for the non-linear relationship in the association model, and the relationship between covariable and PTLD was characterized by a global p-value and by reporting a set of HRs and 95% confidence intervals, one for each component of the piecewise linear spline.

Multiple imputation was used to impute missing values for covariables with at most 25% missingness. Specifically, missing data were filled in ten times using the Full Condition Specification method to generate ten complete data sets, the ten complete data sets were analyzed using standard statistical analyses, and the results from the ten complete data sets are combined using Rubin’s Rule to produce the final association results. Multiple imputation was performed assuming that the data are missing at random ^23^. Continuous, binary, and discrete variables were imputed using linear regression, logistic regression, and the discriminant function, respectively, using observed ranges as bounds on the imputed values.

For both age and EBV match, the HR was allowed to vary continuously with event time (linearly on the log-hazard scale). This was achieved by creating an interaction term between each of the covariates and event time. As such, association results for age and EBV match are listed under the time-dependent covariables section of table 3. Additionally, association between age and time to PTLD was found to be nonlinear. To account for this nonlinearity, a linear spline with 2 knots at baseline age of 45 and 62 years was used. This approximation models the relationship between age and the hazard of PTLD linearly, but it allows the slope of the linear relationship to vary over 3 ranges created by the knots. Thus, a HR for age less than 45 years, between 45 and 62 years, and greater than 62 years are reported. Each HR describes the multiplicative change in the hazard of PTLD for a 5-year change in recipient age. This approximation of the non-linear relationship and the knots were chosen based on the RCS spline fit which allows more complex non-linear relationship.

**Table 2.** Cumulative event counts and Kaplan Meier rates for PTLD.

|  | **Cumulative**<br>**Event Count<sup>+</sup>** | **Number of**<br>**Patients at Risk** | **KM Event**<br>**Probability (95% CI)<sup>\*</sup>** |
| --- | --- | --- | --- |
| **At 1 Year** | 215 | 17328 | 1.14% (0.99%, 1.29%) |
| **At 3 Years** | 319 | 11150 | 1.85% (1.65%, 2.05%) |
| **At 5 Years** | 376 | 6740 | 2.47% (2.21%, 2.73%) |
| **At 10 Years** | 436 | 1588 | 4.12% (3.60%, 4.63%) |
| **At 15 years** | 453 | 143 | 6.54% (5.12%, 7.96%) |
| + A total of 454 PTLD events were observed in the analysis cohort with a maximum follow-up time of 17 years. |  |  |  |
| * The Kaplan Meier method estimates the probability an event occurs on or before a given time point. |  |  |  |

**Table 3.** Association between pre- and peri-transplant characteristics and time-to-PTLD development.

|  | Univariable Models |  | Multivariable Model |  |
| --- | --- | --- | --- | --- |
|  | HR (95% CI) | P-value | HR (95% CI) | P-value |
| **Time Dependent Covariates** |  |  |  |  |
| Age (Per 5 Years) |  | *<0.0001* |  | *<0.0001* |
| At Year 1 | - | - | - | - |
| Age Less than 45 Years | 0.69 (0.63, 0.77) | - | 0.79 (0.70, 0.89) | - |
| Age Between 45 and 62 Years | 1.39 (1.23, 1.58) | - | 1.42 (1.25, 1.62) | - |
| Age Greater than 62 Years | 0.73 (0.56, 0.96) | - | 0.70 (0.54, 0.93) | - |
| At Year 5 | - | - | - | - |
| Age Less than 45 Years | 0.80 (0.71, 0.91) | - | 0.89 (0.78, 1.02) | - |
| Age Between 45 and 62 Years | 1.31 (1.15, 1.49) | - | 1.34 (1.18, 1.54) | - |
| Age Greater than 62 Years | 0.81 (0.51, 1.29) | - | 0.77 (0.48, 1.23) | - |
| At Year 10 | - | - | - | - |
| Age Less than 45 Years | 0.96 (0.75, 1.24) | - | 1.03 (0.80, 1.34) | - |
| Age Between 45 and 62 Years | 1.21 (0.93, 1.57) | - | 1.25 (0.97, 1.63) | - |
| Age Greater than 62 Years | 0.92 (0.33, 2.58) | - | 0.85 (0.30, 2.41) | - |
| EBV Match |  | *<0.0001* |  | *<0.0001* |
| Donor Negative/Recipient Negative | - | - | - | - |
| At Year 1 | 4.93 (2.47, 9.82) | - | 4.65 (2.34, 9.23) | - |
| At Year 5 | 3.04 (1.20, 7.69) | - | 3.00 (1.22, 7.38) | - |
| At Year 10 | 1.66 (0.26, 10.8) | - | 1.74 (0.28, 10.7) | - |
| Donor Positive/Recipient Negative | - | - | - | - |
| At Year 1 | 5.15 (3.96, 6.69) | - | 5.00 (3.81, 6.56) | - |
| At Year 5 | 2.55 (1.64, 3.97) | - | 2.60 (1.69, 4.00) | - |
| At Year 10 | 1.06 (0.38, 2.99) | - | 1.15 (0.42, 3.16) | - |
| Recipient Positive | - | - | - | - |
| **Time Independent Covariates** |  |  |  |  |
| Native Lung Disease |  | *0.0056* |  | *0.0014* |
| Vascular | 1.19 (0.69, 2.06) | - | 1.37 (0.78, 2.43) | - |
| Cystic Fibrosis | 1.81 (1.39, 2.35) | - | 1.47 (0.95, 2.26) | - |
| Restrictive | 1.47 (1.19, 1.83) | - | 1.57 (1.25, 1.96) | - |
| Obstructive | - | - | - | - |
| Bilateral Transplant Type | 1.09 (0.90, 1.33) | 0.3657 | 1.09 (0.86, 1.38) | 0.4803 |
|  | HR (95% CI) | P-value | HR (95% CI) | P-value |
| Male | 1.30 (1.08, 1.58) | 0.0059 | 1.17 (0.96, 1.43) | 0.1212 |
| Ischemia Time | 1.03 (0.97, 1.09) | 0.3270 | 1.03 (0.97, 1.10) | 0.3605 |
| ABO Exact Match | 1.10 (0.77, 1.57) | 0.6126 | 1.11 (0.77, 1.59) | 0.5783 |
| Functional Status at Time of Transplant |  | 0.1434 |  | 0.2641 |
| Performs Activities of Daily Living with Total Assistance | 0.94 (0.70, 1.28) | - | 0.91 (0.67, 1.24) | - |
| Performs Activities of Daily Living with Some Assistance | 0.80 (0.62, 1.03) | - | 0.81 (0.63, 1.05) | - |
| Performs Activities of Daily Living with No Assistance | - | - | - | - |
| Serum Albumin Prior to Transplant | 0.88 (0.73, 1.06) | 0.1701 | 1.02 (0.83, 1.26) | 0.8219 |
| History of Prior Malignancy | 0.91 (0.60, 1.37) | 0.6516 | 0.96 (0.63, 1.46) | 0.8496 |
| PRA% Class I > 0 | 0.85 (0.64, 1.14) | 0.2685 | 0.91 (0.67, 1.24) | 0.5404 |
| PRA% Class II > 0 | 0.80 (0.56, 1.13) | 0.1949 | 0.87 (0.60, 1.26) | 0.4613 |
| Donor/Recipient CMV Match |  | 0.0045 |  | 0.2133 |
| Donor Negative/Recipient Negative | 1.41 (1.10, 1.81) | - | 1.21 (0.93, 1.56) | - |
| Donor Positive/Recipient Negative | 1.36 (1.08, 1.71) | - | 1.19 (0.94, 1.51) | - |
| Recipient Positive | - | - | - | - |
| Recipient HCV Positive | 1.01 (0.48, 2.12) | 0.9759 | 1.10 (0.52, 2.30) | 0.8075 |
| Donor History of High Risk Behaviors | 1.42 (1.01, 2.01) | 0.0459 | 1.42 (1.01, 2.01) | 0.0489 |
| Donor Age (Per 5 Years) | 0.97 (0.94, 1.00) | 0.0917 | 0.98 (0.95, 1.02) | 0.3760 |
| Donor History of Cancer | 0.53 (0.10, 2.77) | 0.4355 | 0.59 (0.11, 3.14) | 0.5209 |
| Donor Steroids | 1.09 (0.85, 1.39) | 0.4888 | 1.10 (0.86, 1.40) | 0.4616 |
| *Univariable = model includes <u>only</u> one covariate; Multivariable = model includes <u>all</u> covariates listed<br>HR = hazard ratio; CI = confidence interval |  |  |  |  |

## RESULTS

### Patient characteristics

The ISHLT Registry data for 49,776 patients receiving lung transplantation between January 2006 and June 2015 were included. Recipients excluded from analysis included: 3852 recipients with less than 1 year of follow up after lung transplantation, 1413 recipients of re-transplantation and/or lobar lung transplant, 24572 patients from centers not reporting PTLD during follow-up and 630 patients younger than 18 years of age (figure 1). Thus, the analysis cohort included 19,309 adult first lung transplant recipients.

**Figure 1:**
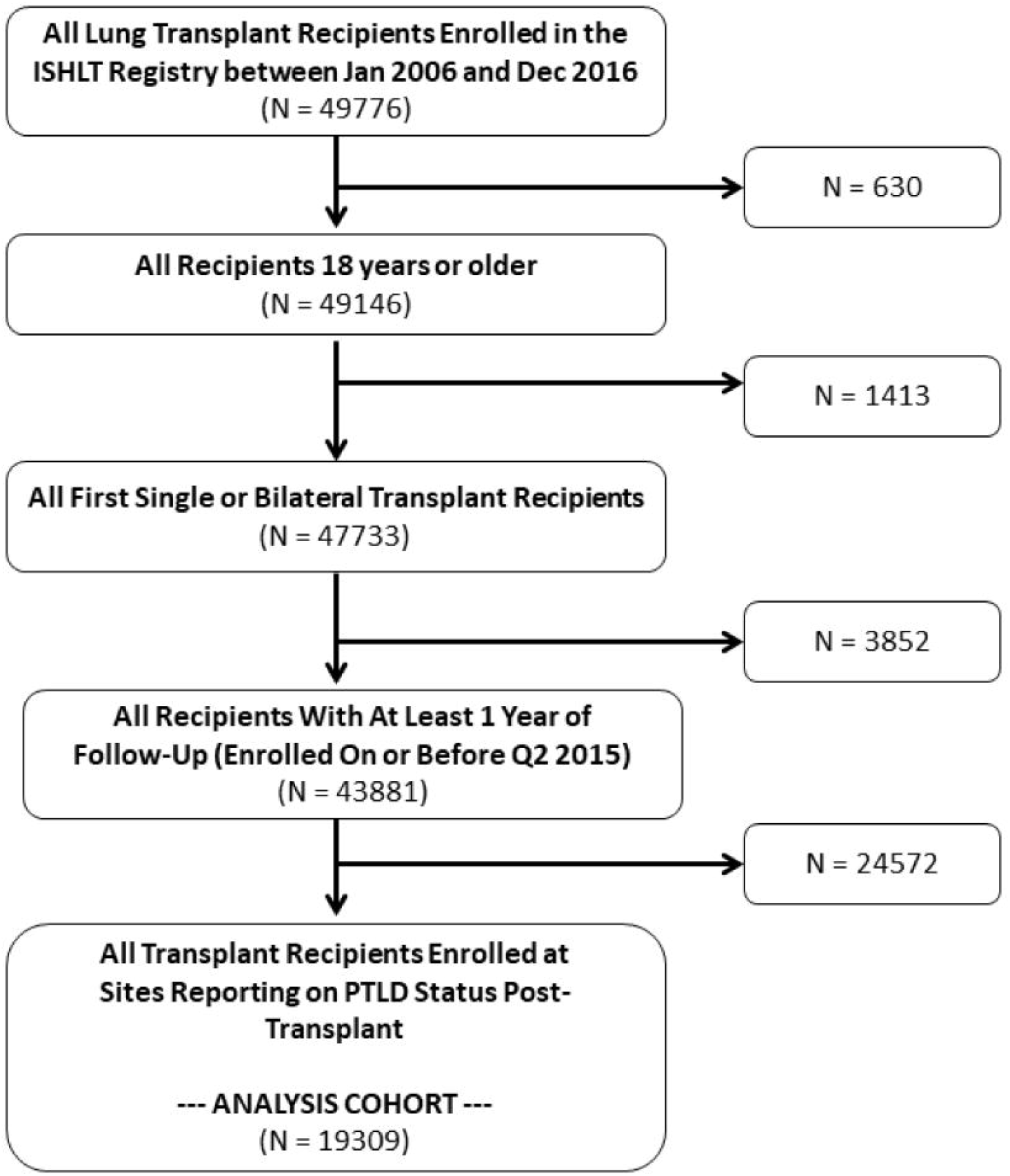
Flowchart showing selection of the final study population.

Median age at transplantation was 57 years and 43.1% were women. Most common indication for transplantation was restrictive disease (43.2%) followed by obstructive disease (39.9%) and cystic fibrosis (13.6%). PTLD occurred in 454 (2.45%) patients after lung transplantation. Clinical characteristics of study cohort are described in table 1. Distribution of these covariates were similar between patients with and patients without PTLD. However, we observed some notable differences. At time of transplant, only 53.3% of recipients who subsequently developed PTLD were EBV seropositive, compared to 77% in recipients who did not develop PTLD. We noted that EBV mismatch (D +/R-) was reported in 18.16% of cases in recipients who subsequently developed PTLD but in only 6% of patients who did not.

### Incidence of PTLD

The cumulative incidence of PTLD was 1.14% (95% CI = 0.99%, 1.29%) during the first year, 2.47%% (95% CI= 2.21%, 2.73%) at 5-years, and 4.12% (95% CI= 3.6%, 4.63%) at 10-years after transplantation (figure 2 and table 2). Of those who developed PTLD, 47.4% (215 out of 454) of cases occurred within 1-year after lung transplantation.

**Figure 2:**
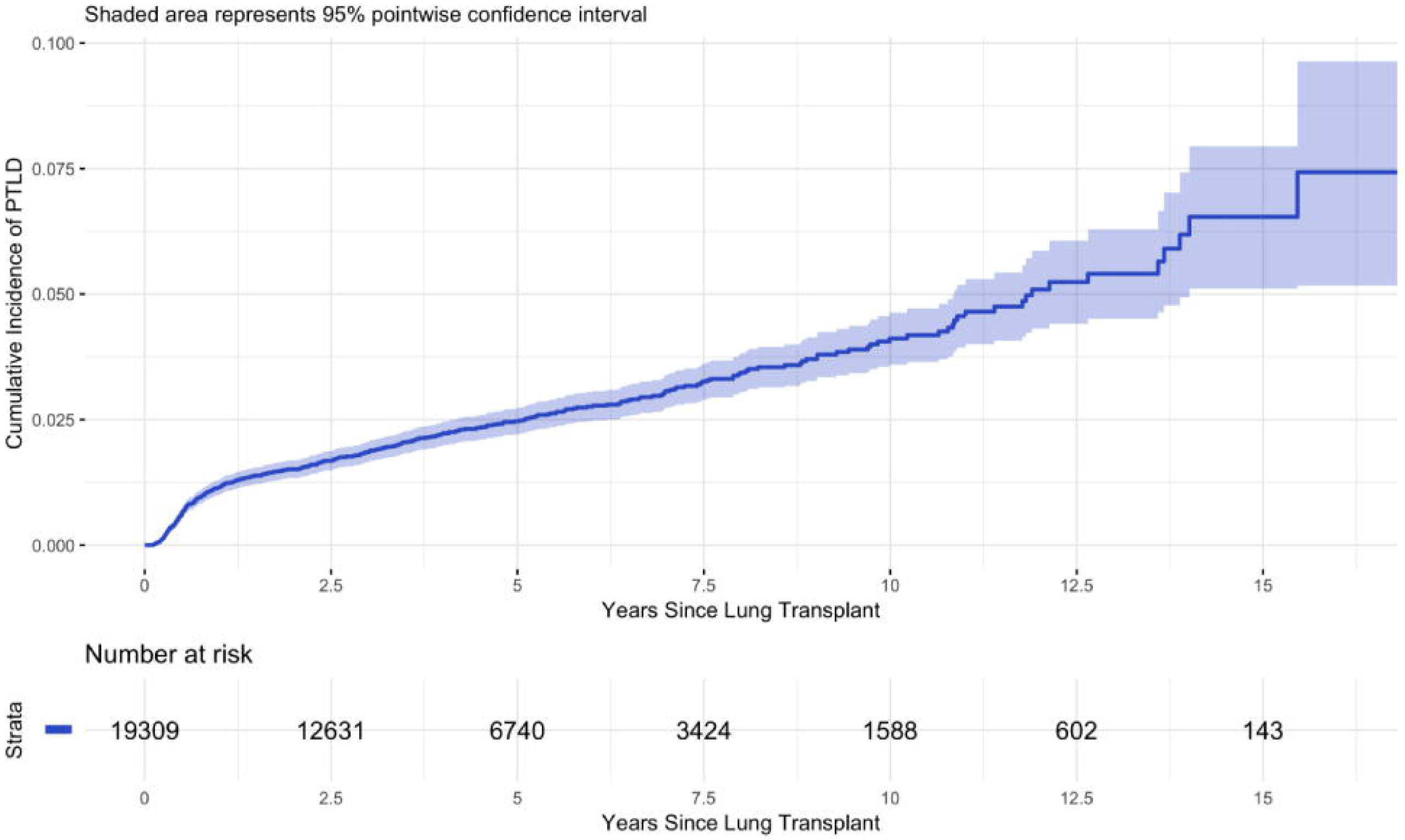
Cumulative incidence of PTLD after lung transplant in a cohort of 19309 lung transplant recipients. The numbers below the curve are numbers of patients being followed up at yearly intervals post-lung transplant and at risk of developing PTLD.

### Risk Factors for PTLD

Results of univariable and multivariable Cox regression modeling for time to PTLD are presented in table 3. When exploring the association between pre- and peri-transplant characteristics and time to PTLD, the proportional hazards assumption was found to be violated for recipient age and EBV matching, suggesting that the association between each of these characteristics and hazard of developing PTLD changes during the follow-up period. To account for this non-proportionality, time-dependent covariates were created for these characteristics, as described in Methods.

Both unadjusted and adjusted associations between age and development of PTLD were found to be statistically significant (p<0.0001 for both models). Using the multivariable model, we found that at 1-year post-transplantation risk of PTLD decreased with increasing age for subjects younger than 45 years at time of transplant (HR 0.69 per 5 years increase; 95% CI, 0.63, 0.77); then the risk of PTLD increased with increasing age for subjects between 45 and 62 years at time of transplant (HR 1.39 per 5 years increase; 95% CI, 1.23, 1.58), and finally risk of PTLD decreased with increasing age for subjects older than 62 years at time of transplant (HR 0.73 per 5 years increase; 95% CI, 0.56, 0.96). A similar pattern was observed at 5 and 10-years post-transplantation. However, the strength of the association diminished over time; remaining statistically significant at 5-years post-transplantation but not at 10-years (figure 3, table 3).

**Figure 3:**
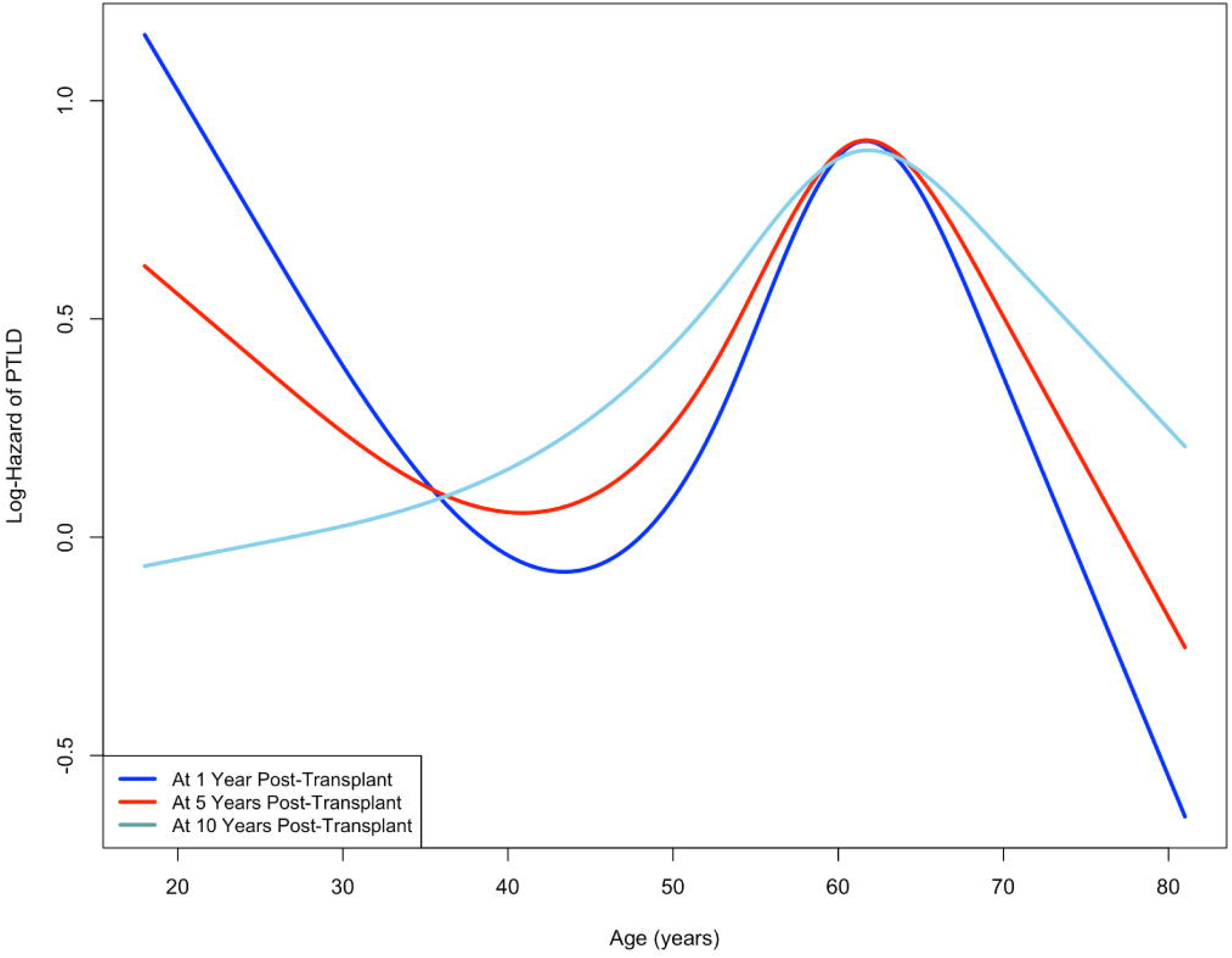
Adjusted log-Hazard of PTLD as function of age at transplant. A linear spline with 2 knots at baseline age of 45 and 62 years was used to account for the nonlinear association between age at transplant and time to development of PTLD. Hazard Ratio (HR) for 3 age groups: less than 45 years old, between 45 and 62 years, and greater than 62 years old are reported.

Similarly, in both unadjusted and adjusted association models, EBV serostatus match between donor and recipient was found to be significantly associated with PTLD (p<0.0001 for both models). Using the multivariable model, we found that at 1-year post-transplantation, relative to a transplant in which recipient was EBV seropositive (regardless of donor status), transplants in which donor and recipient were EBV status negative (D-/R-) (HR 4.93; 95% CI, 2.47, 9.82) and mismatched transplants (D+/R-) (HR 5.15; 95% CI, 3.96, 6.69, p <0.0001) were associated with an increased risk of PTLD. The strength of both of these associations attenuated over time; remaining statistically significant at 5-years but not at 10-years post-transplantation.

We then analyzed the association between time-independent covariables and PTLD after lung transplantation. We observed that native lung disease (p = 0.0056) and donor history of high risk behaviors (p = 0.04) were found to be independently associated with PTLD in the multivariable model. In adjusted association models, recipients with CF (HR 1.81; 95% CI, 1.39, 2.35) and restrictive disease (HR 1.47; 95% CI, 1.19, 1.83) had a statistically significant increased risk of PTLD compared to those with obstructive disease. Transplant recipients whose donors had a history of high risk behaviors had a statistically significant increased risk of PTLD (HR 1.42; 95% CI 1.01, 2.01). In the unadjusted model, it was found that males had a higher risk of PTLD compared to females (HR 1.30; 95% CI, 1.08, 1.58, p = 0.006). Although the same direction of association was found after adjusting for the other characteristics, the adjusted association was not found to be statistically significant (p = 0.10). Finally, in the unadjusted model, relative to CMV seropositive recipients (regardless of donor status), transplants in which donor and recipient were CMV negative (D-/R-) (HR 1.41; 95% CI, 1.1,1.81) and mismatched transplants (D+/R-) (HR 1.36; 95% CI, ,1.71, p 0.0045) were associated with an increased risk of PTLD. The adjusted association was not found to be statistically significant (p=0.21)

### HLA and risk of PTLD

We analyzed the unadjusted and adjusted association between donor and recipient HLA specificities and risk of PTLD. Serological HLA typing was reported for donors and recipients to the ISHLT Registry. Transplants involving donors with HLA-A1 were associated in both models with an increased risk of PTLD compared to transplants involving donors without HLA-A1 (HR 1.26; 95% CI, 1.03, 1.56, p = 0.03 in the adjusted model). Similarly, transplants involving donors with HLA-A24 were associated with an increased risk of PTLD compared to transplants involving donors without HLA-A24 in both models (HR 1.30; 95% CI, 1.03, 1.64, p = 0.03 in the adjusted model). Transplants involving recipients with HLA-DR11 were associated with a decreased risk of PTLD compared to transplants involving donors without HLA-DR11 in both models (HR 0.74; 95% CI, 0.55, 0.99, p =0.045 in the adjusted model). (Supplemental Table 1 and Figure 4).

**Figure 4:**
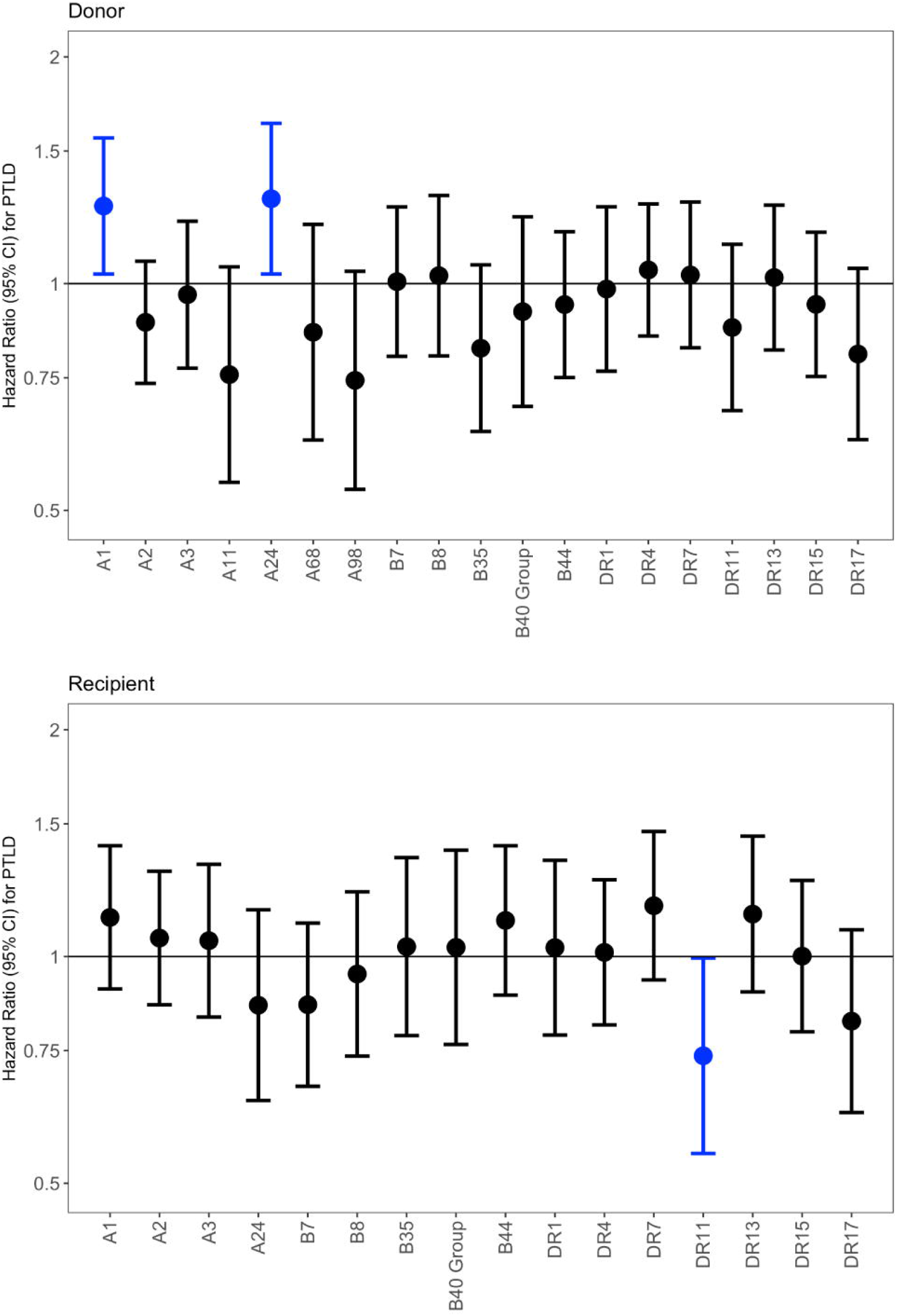
Hazard ratio for PTLD by HLA type Donors (top); Recipients (bottom).

## DISCUSSION

PTLD is a rare but life-threatening complication following transplantation. Several single-center analyses have identified numerous risk factors associated with PTLD such as primary EBV infection, type of organ transplanted, young age, Caucasian race, and CMV mismatch. Older age is considered a risk factor for PTLD after the first post-transplant year ^13,15,24^. However, these analyses often present limitations due to their small size ^1,8,24^. For the first time with the present study, we aimed to evaluate the incidence and identify clinical risk factors associated with development of PTLD in a large, international multi-center adult lung transplant recipient population using the ISHLT Registry. The most important findings of this study are that despite low overall incidence, we noted that the cumulative incidence reaches 4.4% at 10-years post-transplantation. We observed a nonlinear association between age and risk of PTLD, with increasing risk in lung transplant recipients between 45-62 at transplant, but decreasing risk with age below and above this age range. Other risk factors were a negative EBV serostatus, CF and restrictive lung disease. Finally, we observed an increased risk of PTLD in recipients of donor organs with HLA-types A1 and A24, and decreased risk in recipients with HLA-DR11.

In this study of the ISHLT Registry database, we found that there is a sustained increase in the cumulative risk of developing PTLD following lung transplant. The overall incidence of PTLD is small, lower than reported in prior studies in lung transplantation^8-10,14,16^. However, several centers participating in the ISHLT Registry have not reported PTLD consistently over time and this might explain lower than expected incidence. Similar to previous studies describing a higher incidence of PTLD within first year after lung transplantation^2,7,13^, we noted that approximately 50% of PTLD cases occurred during the first year after lung transplant. Several factors could explain this heightened risk, including sudden appearance of an EBV-positive organ in a naïve recipient (in EBV mismatch transplantation) and intensity of immunosuppression. Particularly during the first year after transplantation, stronger immunosuppression and less efficient host immune responses against EBV might predispose to malignant transformation of EBV-infected cells.

Age is an important risk factor for PTLD at different time after transplantation. Younger individuals have a high risk of PTLD during first year, while older recipient age has emerged as a risk factor for PTLD after the first year from transplant ^15^. We observed a similar risk in lung transplant recipients. In our detailed analysis around recipient age, we noted a non-linear association between age and risk of PTLD in adult lung transplant patients. Risk of PTLD declined with increasing age for recipients younger than 45 years, while risk of PTLD increased for recipients between 45-62 years at time of transplant. Retrospective analyses of kidney and heart transplant recipients reported an increased risk of PTLD in patients older than 50 years after the first year following transplantation ^25,26^.

Our study confirms that lung transplant recipients with negative EBV serostatus, and in particular EBV mismatch recipients, have an increased risk of PTLD, suggesting that this specific population requires more attentive monitoring following transplantation. The role of CMV in development of PTLD remains contradictory. Initial single-center experiences reporting increased risk of PTLD in CMV mismatch SOT recipients ^27^have not been confirmed in thoracic transplant recipients ^9 13^. In our analysis, CMV negative recipient status was only associated with PTLD in the unadjusted model, confirming that CMV mismatch or infection should not be considered a major risk factor in lung transplant recipients

We observed that lung transplant recipients with CF and restrictive disease have an increased risk of PTLD compared to other lung diseases. These findings are partially explained by the EBV serostatus and age of these two populations; lung transplant recipients with CF are more often young and EBV seronegative, while patients with restrictive disease tend to be older. Although several possible explanations for this finding could be considered, EBV infection has been implicated in the pathogenesis of restrictive lung disease implying loss of immune surveillance in this patient group ^28^.

We investigated the association between HLA and PTLD. Previous observational reports demonstrated increased risk of PTLD with different HLA alleles. European studies in SOT recipients suggested that HLA-A18, B21, B45 and HLA-DR13 were associated with increased risk of PTLD ^18,20^. Two single-center analyses from U.S. centers described increased risk of PTLD with HLA A-26, B8 and B40 group and risk reduction associated with donor HLA-A1, B8 and DR3 ^19,21^. These associations are different from those found in our international Registry analysis, perhaps because of genetic differences between the populations studied. We found that recipients with HLA-DR11 have a decreased risk of PTLD, while donor haplotype A1 and A24 are associated with increased risk of PTLD. However, our conclusions should be considered in light of several limitations. We were limited to including only serological HLA typing, rather than molecular typing, which could provide a more detailed assessment of any association. Moreover, we could not analyze the impact of donor/recipient HLA mismatching. However, our analysis included a larger cohort compared to previous analyses and serological typing was also used in the majority of those studies.

This study presents several further limitations which should be acknowledged. This was a retrospective analysis that linked pre- and peri-transplant clinical characteristics with subsequent PTLD reporting. Thus, we cannot draw any conclusions regarding causation. Of note, the ISHLT Registry was not specifically developed to look at PTLD and therefore this study is limited to the data captured by the Registry. Furthermore, some variables (e.g., rejection and increased immunosuppression) had a large percentage of missing data or was not captured in the Registry, and therefore could not be included in the analysis. This potentially limits both generalizability of study’s results and broader evaluation of incidence and risk factors.

In conclusion, this analysis describes a large cohort of adult lung transplant recipients with PTLD, allowing meaningful assessment of incidence and risk factors. We noted an increasing cumulative incidence of PTLD following lung transplantation, with the highest risk during first year. EBV seronegative recipients were at highest risk of PTLD, regardless of donor serostatus and independent of other factors, compared to seropositive recipients. We noted a complex relationship between age at transplant and PTLD risk, and an increased risk, independent of other factors, in recipients with CF and restrictive lung disease. Our findings will assist clinicians caring for patients after lung transplantation to target PTLD surveillance to the populations at highest risk.

## Data Availability

The authors confirm that the data supporting the findings of this study are available within the article and its supplementary materials.

## Notes

### Competing Interest Statement

The authors have declared no competing interest.

### Funding Statement

This work was supported by the International Heart and Lung Transplant Registry through the Transplant Registry Early Career Award

